# Revisiting the DIGIT-HF Trial: Evidence of Time-Dependent Treatment Effects

**DOI:** 10.1101/2025.11.22.25339857

**Authors:** Aldama-López Guillermo, López-Vázquez Domingo, Rebollal-Leal Fernando

**Affiliations:** Cardiology Department, University Hospital of A Coruña, Spain; Institute for Biomedical Research of A Coruña (INIBIC), Spain

**Keywords:** Heart failure, digitoxin, proportional hazards, time-dependent treatment effect, survival analysis, Cox regression

## Abstract

**Background:** The DIGIT-HF trial reported that digitoxin reduced the composite risk of all-cause mortality or first heart failure hospitalization in patients with heart failure and reduced ejection fraction, summarized by a single hazard ratio (HR) of 0.82. The validity of this single estimand depends on the proportional hazards (PH) assumption of the Cox model.

**Methods:** We performed a methodological re-analysis based on individual patient data reconstructed from the published Kaplan-Meier curves of the DIGIT-HF trial’s primary endpoint. We formally tested the PH assumption, modeled the time-dependent treatment effect, and performed a pre-specified landmark analysis at 18 months.

**Results:** Our analysis confirmed a statistically significant violation of the proportional hazards assumption for the overall follow-up period (p = 0.019). The time-dependent HR plot showed that the beneficial effect of digitoxin waned over time. The landmark analysis revealed a significant benefit in the first 18 months (HR, 0.69; 95% CI, 0.54–0.88) but no evidence of benefit thereafter (HR, 0.99; 95% CI, 0.77–1.28). The PH assumption was met within both the early (p = 0.17) and late (p = 0.54) periods.

**Conclusion:** Out results strongly suggest the treatment effect of digitoxin in DIGIT-HF is not constant over time, invalidating use of a single hazard ratio. Benefit is confined to the first 18 months of therapy, challenging the interpretation of digitoxin as providing sustained benefit. These findings have important implications for clinical practice and highlight the necessity of testing proportional hazards assumptions in cardiovascular trials.

## Introduction

The Digitoxin to Improve Outcomes in Chronic Heart Failure (DIGIT-HF) trial concluded that in patients with heart failure with reduced ejection fraction (HFrEF), digitoxin reduced the composite primary outcome of all-cause mortality or first heart failure hospitalization, reporting a hazard ratio (HR) of 0.82 (95% CI, 0.69–0.98).^1^ This result was derived from a Cox proportional hazards model. However, visual inspection of the published cumulative incidence curves suggested potential violation of the proportional hazards (PH) assumption—a fundamental prerequisite for the validity of a single, time-averaged HR.^2^ When hazards are not proportional over time, the reported constant HR becomes a misleading summary that may obscure clinically important temporal patterns in treatment effect. We performed a methodological re-analysis to formally investigate the PH assumption and characterize the temporal profile of digitoxin’s treatment effect.

## Methods

We reconstructed individual patient data from published Kaplan-Meier curves using the validated algorithm by Guyot et al.,^3^. To validate our reconstruction, we fitted a Cox model to comparing it with the original publication.

We assessed the PH assumption using three complementary approaches. First, we generated log-log survival curves, which should be parallel under the PH assumption. Second, we conducted formal statistical testing using scaled Schoenfeld residuals, where a significant p-value indicates rejection of the PH assumption.^4^ We also plotted smoothed scaled residuals against follow-up time to visualize potential time-dependent patterns in the log hazard ratio. Third, we fitted a flexible parametric survival Royston-Parmar model using restricted cubic splines to estimate the time-varying HR as a continuous function of time.^5^

The time-dependent HR model revealed that the upper limit of the 95% confidence interval crossed 1.0 at approximately 16 months. Based on this finding, and rounding to a clinically interpretable time point, we performed a landmark analysis at 18 months to formally quantify treatment effects in early versus late periods. For the early period (0–18 months), patients with events beyond 18 months were censored at the landmark time. For the late period (>18 months), only patients’ event-free at 18 months were included, with time recalculated from the landmark point. The PH assumption was tested within each period.

All analyses were performed in R version 4.2.2. A p-value <0.05 was considered statistically significant.

## Results

Our reconstructed data replicated the primary results exactly (HR 0.82; 95% CI, 0.69–0.98), confirming validity of our reconstruction.

The log-log survival curves were non-parallel (Figure, panel A), and scaled Schoenfeld residuals showed a significant time trend with increasing values over follow-up (Figure, panel B), indicating the log hazard ratio changed systematically over time. Formal assessment revealed significant violation of the PH assumption (p=0.019).

Time-dependent modeling demonstrated that digitoxin’s benefit waned progressively throughout follow-up (Figure, panel C). This contrasts sharply with the constant HR of 0.82 reported in the trial, which assumes uniform treatment effect throughout follow-up.

The landmark analysis at 18 months quantified this temporal heterogeneity precisely (Figure, panel D). In the first 18 months, digitoxin provided substantial benefit (HR 0.69; 95% CI, 0.54–0.88; p<0.001). Beyond 18 months, there was no evidence of benefit (HR 0.99; 95% CI, 0.77–1.28; p=0.94). Critically, the PH assumption was satisfied within both early (p=0.17) and late (p=0.54) periods, confirming this stratified analysis provides valid representation of treatment effects in each timeframe.

## Discussion

Our analysis demonstrates that digitoxin’s treatment effect in DIGIT-HF is not constant over time. The reported single HR of 0.82 masks complete absence of benefit beyond 18 months—a finding with important clinical implications for contemporary heart failure management.

Time-varying treatment effects are increasingly recognized in cardiovascular trials. The ISCHEMIA trial, comparing invasive versus conservative management of stable coronary disease, showed crossing hazards with early procedural harm followed by late benefit.^6^ In that trial, the overall HR of 0.93 masked fundamentally different effects across time periods. Similarly, our analysis reveals digitoxin’s benefit is time limited.

Several biological mechanisms could explain this pattern. Tachyphylaxis or receptor desensitization may develop with chronic digitalis exposure, reducing positive inotropic and neurohormonal effects over time. Progressive ventricular remodeling may eventually overwhelm digitoxin’s beneficial effects on myocardial energetics. Competing risks, particularly non-cardiovascular mortality, may become more prominent with longer follow-up. Treatment adherence may decline, or clinicians may have adjusted doses in response to side effects. Unfortunately, reconstructed data preclude assessment of these hypotheses.

The clinical implications are substantial. First, digitoxin may be most appropriate for time-limited use rather than indefinite therapy in HFrEF. Second, the risk-benefit profile warrants reassessment beyond 18 months, particularly given digitoxin’s narrow therapeutic window and toxicity potential. Third, treatment de-escalation or discontinuation strategies warrant prospective investigation in patients remaining stable after initial therapy.

Our findings underscore broader methodological concerns. Nonproportional hazards are common in cardiovascular trials but frequently undetected.^7^ When present, a single HRs can profoundly mislead. Alternative approaches provide more interpretable summaries if treatment effects vary temporally.^8^

Importantly, the DIGIT-HF investigators did not report testing the PH assumption. This reflects a broader problem in cardiovascular trial reporting. Journals should mandate formal PH testing and pre-specified analytic plans for handling violations.

While IPD reconstruction is validated,^3^ limitations exist. We cannot assess adherence patterns, competing risks in detail, differential dropout, or treatment effect heterogeneity across subgroups defined by baseline characteristics. Access to original trial data would enable more detailed analyses.

Nevertheless, our reconstruction’s exact replication of published results strongly supports validity of our conclusions. The statistical evidence for nonproportional hazards is robust across multiple analytical approaches.

## Conclusion

The DIGIT-HF trial’s primary analysis, based on a single time-averaged HR, masks clinically important temporal variation in treatment effect. Rigorous re-analysis strongly suggest digitoxin’s benefit is concentrated in the first 18 months, with no evidence of sustained efficacy thereafter. This calls into question the interpretation that digitoxin provides lasting benefits and suggests the need for limited-duration treatment strategies in clinical practice. More broadly, our analysis underscores the critical importance of verifying proportional hazards assumptions in heart failure trials, as violations can profoundly mislead clinical interpretation and practice.

## Data Availability

Complete code and reconstructed data are publicly available at https://github.com/galdlop/digit_hf/tree/main.

## Funding

None.

## Conflict of Interest

None declared.

---

**Figure:**
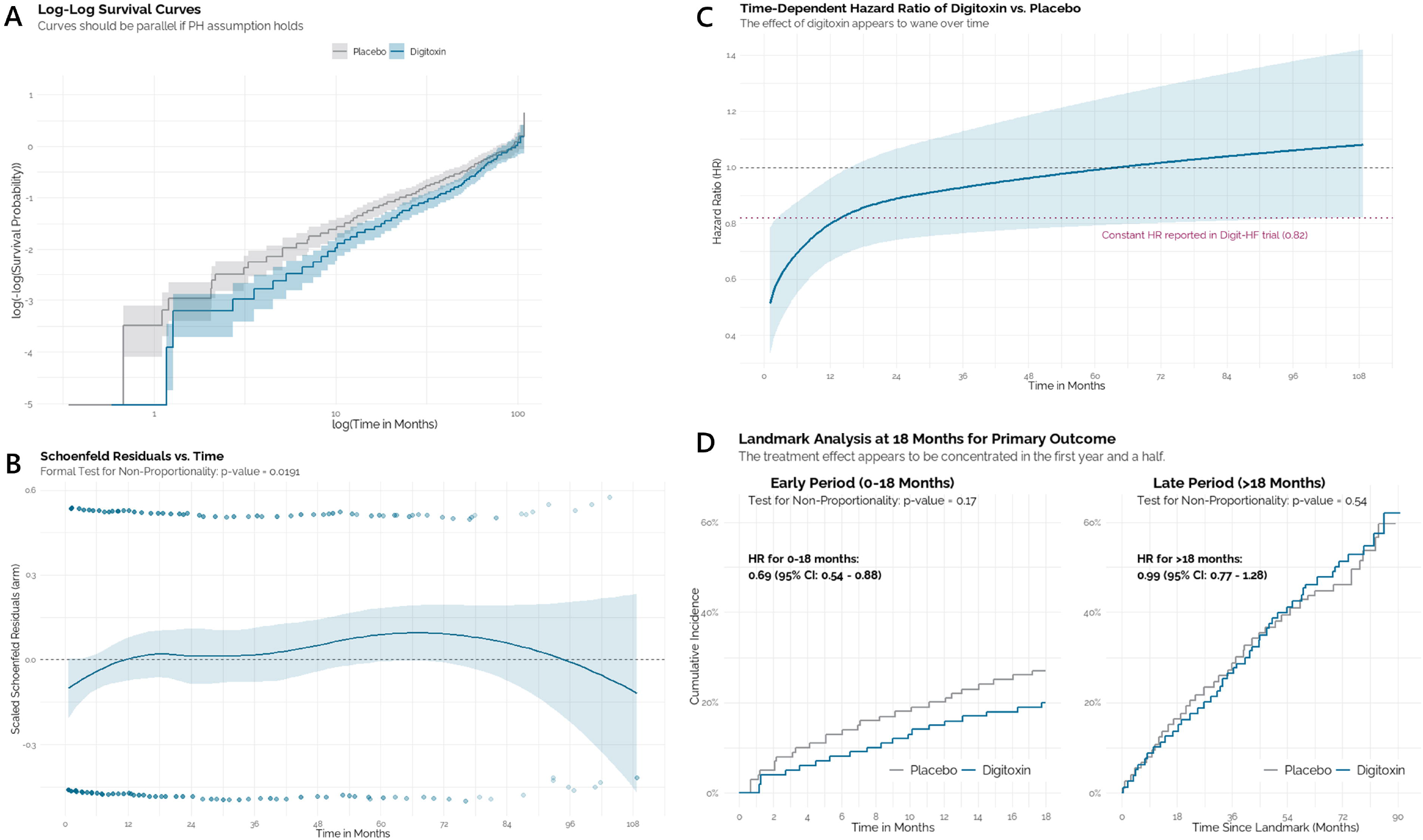
A) Log-Log Survival Curves. The non-parallel nature of the curves for the placebo and digitoxin arms provides initial visual evidence against the proportional hazard’s assumption. **B) Scaled Schoenfeld Residuals Plot**. The plot shows a significant non-zero slope over time, confirmed by the formal test for non-proportionality (p = 0.019). **C) Time-Dependent Hazard Ratio** of Digitoxin vs. Placebo. The solid blue line shows the HR changing over time, with the benefit waning and crossing the null effect line around 18 months. The dotted red line represents the constant HR of 0.82 reported in the trial. **D) Landmark Analysis at 18 Months**. (Left Panel) Analysis of the early period (0-18 months) shows a significant treatment benefit (HR 0.69); the PH assumption is met (p=0.17). (Right Panel) Analysis of the late period (>18 months) shows no evidence of benefit (HR 0.99); the PH assumption is met (p=0.54).

## Notes

### Competing Interest Statement

The authors have declared no competing interest.

